# Bipolar Local Impedance Delta as a Quantitative Index of Catheter–Tissue Energy Coupling During Pulsed-Field Ablation

**DOI:** 10.64898/2026.06.30.26356980

**Authors:** Masaomi Kimura, Mei Hiyama, Shogo Hamaura, Yuichi Toyama, Yuji Ishida, Taihei Itoh, Shingo Sasaki, Hirofumi Tomita

## Abstract

**Background:** Pulsed-field ablation (PFA) systems increasingly provide impedance-based contact indicators, such as tissue proximity indication (TPI), derived from local impedance changes relative to a blood-pool baseline. These indicators are largely binary and do not quantify post-application catheter–tissue energy coupling. We evaluated Bipolar Local Impedance Delta (BiLID), the peri-application bipolar local impedance drop, as a complementary impedance-based index of delivered energy coupling.

**Methods:** We retrospectively analyzed 1,556 VARIPULSE applications in 23 patients undergoing pulmonary vein isolation. BiLID was derived from 29,822 paired pre-/post-ablation impedance measurements obtained from numeric local impedance readouts displayed by the mapping system, without proprietary data export or waveform estimation. Reproducibility was assessed by intraclass correlation. Associations with TPI status, vein anatomy, application order, and peak creatine kinase-MB (CK-MB) were examined using linear mixed-effects models with within-patient clustering and exploratory patient-level analyses.

**Results:** BiLID showed excellent interobserver reproducibility and increased stepwise with the number of TPI-positive electrodes per pair (0–2; P < 0.0001), while varying widely among TPI-positive signals. BiLID differed by electrode position and was lower during right than left pulmonary vein ablation (both P < 0.0001). Total BiLID correlated with peak CK-MB (r = 0.71; 95% CI, 0.42–0.87; P = 0.0001), whereas application count (r = 0.16, P = 0.4711) and TPI-positive signals (r = 0.26, P = 0.2334) did not. Lower CK-MB elevation was associated with larger left atrial volume index, female sex, and heart failure, suggesting substrate-modulated biomarker release.

**Conclusions:** BiLID is a reproducible, continuous index of catheter–tissue energy coupling that complements pre-delivery binary contact indicators by quantifying the response after PFA delivery. It captures graded contact quality and anatomical heterogeneity and may inform individualized, coupling-guided PFA titration.

**Clinical Perspective:** *What is Known:*

- Pulsed-field ablation procedures currently rely largely on cumulative application counts and binary contact-assessment tools such as tissue proximity indication to guide lesion delivery.
- Binary proximity indicators provide useful information on catheter–tissue contact but do not quantify the quality or efficiency of energy coupling during each application.

*What the Study Adds:*

- Bipolar Local Impedance Delta (BiLID), an offline-derived continuous index based on peri-application bipolar local impedance drop, increased stepwise with the number of TPI-positive electrodes and captured variation not represented by binary TPI alone.
- BiLID showed systematic anatomical heterogeneity, with lower values during right pulmonary vein ablation, and correlated with peak CK-MB more strongly than application count or TPI-positive signals.
- Systemic biomarker release was influenced by atrial substrate factors, suggesting that procedural coupling metrics and biomarkers provide complementary information relevant to future individualized PFA titration.

## Introduction

Pulsed-field ablation (PFA) has rapidly emerged as a nonthermal energy source for the treatment of atrial fibrillation. Unlike radiofrequency or cryoballoon ablation, PFA uses ultrarapid high-voltage electrical pulses to induce irreversible electroporation, preferentially affecting cardiomyocytes while sparing adjacent structures.^1^ Large clinical studies have confirmed high procedural success and a favorable safety profile, demonstrating durable pulmonary vein isolation (PVI) with a low incidence of major complications.^2-4^ As adoption widens, attention is shifting from whether PFA can achieve effective PVI toward how the quality of each individual energy delivery can be assessed and optimized.

In contrast to radiofrequency ablation, which relies on established intraprocedural surrogates such as electrode temperature and impedance drop, contemporary PFA systems provide limited quantitative feedback on catheter–tissue interaction during energy delivery. Current guidance depends largely on cumulative application counts or contact-assessment tools derived from unipolar impedance shifts relative to the blood-pool baseline, such as tissue proximity indication (TPI).^5,6^ TPI is reported as a binary signal of contact present or absent, triggered when unipolar impedance rises above a variable baseline. Thus, it provides useful information on tissue proximity but does not quantify the quality or efficiency of catheter–tissue energy coupling. Fluctuations in blood-pool impedance and heterogeneous atrial substrates may further blunt or exaggerate this response, leading to under- or overestimation of effective contact. Moreover, as a binary unipolar metric, TPI cannot characterize the localized biophysical interaction across bipolar electrode pairs during the actual pulse delivery.

The limitations of cumulative metrics are also increasingly relevant. Foundational evidence from the InspIRE trial indicates that a higher number of applications, specifically at least 48, is associated with improved 12-month durability.^2,7^ However, application counts are purely additive and do not account for the quality of each individual delivery. A given number of applications may therefore represent markedly different degrees of effective energy coupling depending on catheter orientation, tissue apposition, regional anatomy, and substrate characteristics.

Preliminary case observations have shown that PFA delivery induces acute local impedance (LI) drops,^8^ suggesting that these shifts may reflect the tissue-level biophysical response to electroporation. To date, however, this phenomenon has been described primarily in qualitative terms, and no standardized continuous index has been established to complement binary proximity information and cumulative application counts.

We therefore propose Bipolar Local Impedance Delta (BiLID), defined as the net LI drop measured immediately before and after each PFA application across bipolar electrode pairs, as a continuous index of catheter–tissue energy coupling derived from peri-application LI changes. By analyzing a comprehensive dataset of individual LI measurements, we investigated whether BiLID provides a graded and reproducible readout that complements binary TPI, whether it captures anatomical heterogeneity in coupling across pulmonary vein regions, and whether it is biologically corroborated by systemic markers of myocardial response. This study aims to provide a quantitative framework for assessing the biophysical quality of individual PFA deliveries and to establish a foundation for future coupling-guided PFA titration.

## Methods

### Study Design and Patient Population

This single-center retrospective observational study included consecutive patients who underwent pulmonary vein isolation (PVI) using pulsed-field ablation (PFA) at Hirosaki University Hospital, Hirosaki, Japan, between October 2025 and January 2026. The inclusion criteria were symptomatic paroxysmal or persistent atrial fibrillation, age 20 years or older, and de novo PVI using a variable-loop PFA system (VARIPULSE; Biosense Webster, Irvine, CA, USA). The exclusion criteria were prior left atrial ablation, additional substrate ablation, and the presence of intracardiac thrombi. The study was approved by the Institutional Review Board of Hirosaki University Hospital (Approval No. 2025-018), and informed consent was obtained using the opt-out methodology. This was an investigator-initiated study. Biosense Webster and Johnson & Johnson had no role in the study design, data collection, analysis, interpretation, or preparation of the manuscript, and provided no financial or material support for this work.

### Ablation Procedure

All procedures were performed under deep sedation using a combination of propofol, thiopental, fentanyl, and morphine, following a previously described protocol.^9^After a single transseptal puncture guided by fluoroscopy and intracardiac echocardiography, a VIZIGO steerable sheath (Biosense Webster) was placed in the left atrium. Unfractionated heparin was administered to maintain an activated clotting time above 350 seconds. Anatomical mapping was performed using an OCTARAY multielectrode mapping catheter (Biosense Webster) and integrated with preprocedural computed tomography images using the CARTO 3 system (Biosense Webster).

PFA was delivered using the TRUPULSE voltage generator (Biosense Webster) and the VARIPULSE variable-loop catheter. The system delivered biphasic bipolar pulse trains at a nominal voltage of 1,800 V. Ablation was performed in sets of three applications per catheter position, with at least four distinct positions targeted per pulmonary vein to ensure circumferential lesion coverage. PVI was confirmed by evaluating both entrance and exit block. When bidirectional block was not achieved, additional PFA applications were delivered at the operator’s discretion for gap closure.

### Local Impedance Monitoring and Definition of BiLID

Real-time local impedance (LI) was monitored using the VARIPULSE PFA system integrated with the CARTO 3 mapping system. Catheter–tissue contact was assessed using tissue proximity indication (TPI), which was displayed as balloon-shaped indicators on the electrode icons within the three-dimensional mapping interface (Figure 1A). LI was measured across 10 adjacent bipolar electrode pairs (B1 through B10) arranged circumferentially on the variable-loop catheter (Figure 1B).

**Figure 1.**
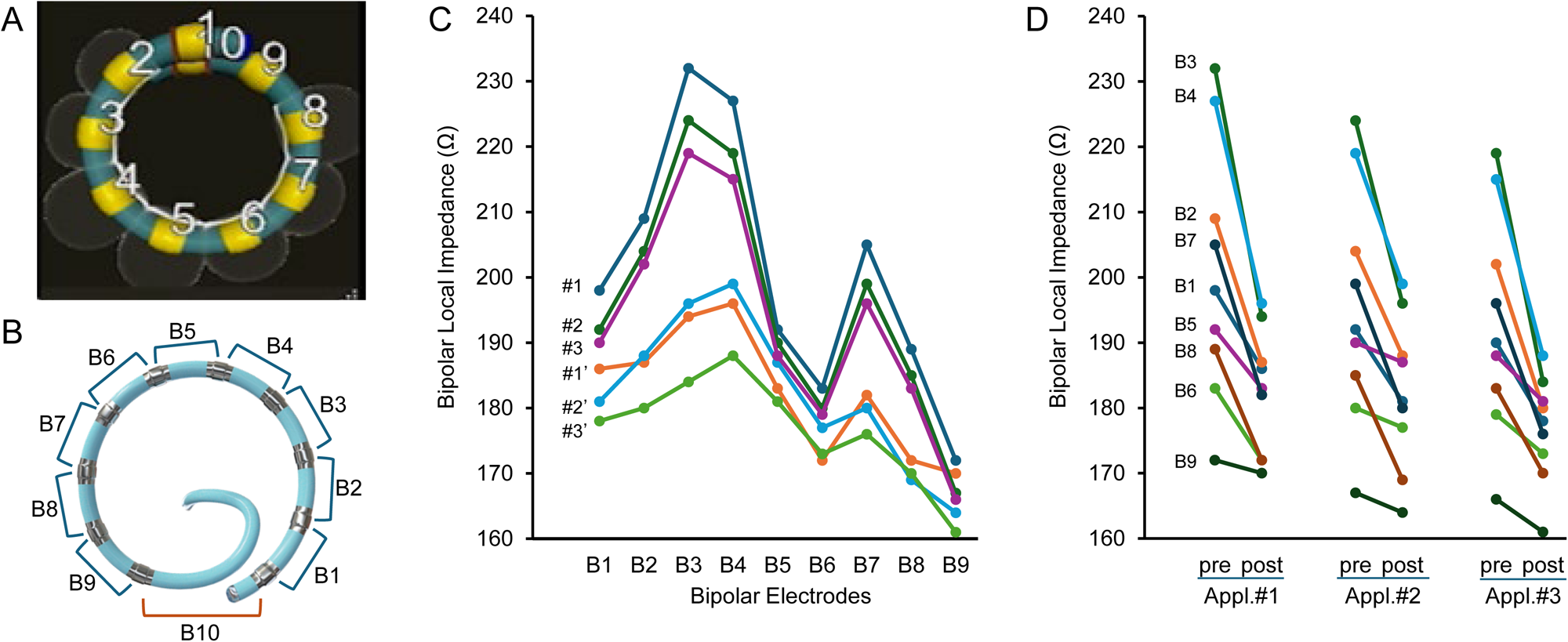
Representative assessment of impedance changes using the variable-loop pulsed-field ablation catheter. A, Representative CARTO three-dimensional mapping display of the variable-loop pulsed-field ablation catheter (VARIPULSE). The numbered electrodes correspond to the loop configuration. Balloon-shaped indicators denote positive tissue proximity indication (TPI) signals on individual electrodes. B, Schematic representation of the VARIPULSE catheter loop, illustrating adjacent bipolar electrode pairs (B1 through B10). Local impedance was monitored across these bipolar pairs during each PFA application. C, Representative line graphs showing local impedance across bipolar pairs during three consecutive PFA applications delivered at the same site. For each application, pre-application and post-application values are shown as paired points. The downward shift from pre-application to post-application values represents the acute impedance drop induced by PFA. D, Slope charts for individual bipolar pairs illustrating the paired pre-to-post application impedance drop for each energy delivery. The magnitude of this paired impedance change defines Bipolar Local Impedance Delta (BiLID). BiLID indicates Bipolar Local Impedance Delta; LI, local impedance; PFA, pulsed-field ablation; and TPI, tissue proximity indication.

BiLID was defined at three hierarchical levels.

At the electrode-pair level, for each bipolar pair i during a given application:

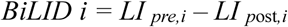

expressed in ohms (Ω).

At the application level, BiLID was summed across all valid bipolar pairs within that application. At the patient level, Total BiLID was the sum of all valid pair-level BiLID values across every application delivered in that patient:

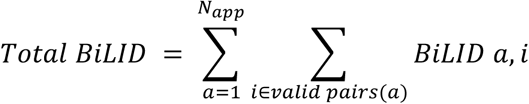

In the present study, BiLID was calculated offline from recorded procedural videos and was not automatically displayed to the operator during the procedure. To ensure precise temporal alignment and minimize measurement drift, the entire procedural user interface was continuously recorded using a high-resolution video capture system. Pre-ablation and post-ablation LI values were systematically extracted by frame-by-frame retrospective analysis of these recordings by an experienced electrophysiologist who was blinded to the patients’ clinical outcomes. Pre-ablation LI was defined as the stable LI value observed in the single frame immediately preceding initiation of the PFA pulse train, and post-ablation LI was measured in the frame immediately following completion of the energy delivery cycle (Figures 1C and 1D). Because BiLID was derived solely from the discrete local impedance values displayed on the system interface, measurement consisted of transcribing these numeric readouts rather than visually estimating values from continuous waveforms or graphs. Accordingly, measurement error was limited to occasional transcription error rather than reader-dependent interpretive variability, and the temporal sampling rate of the video capture did not affect the recorded values.

For cumulative patient-level analysis, Total BiLID was defined as the sum of all valid BiLID values obtained across all delivered energy applications. The number of TPI-positive signals was defined as the patient-level sum of all electrode-level TPI-positive signals observed across completed applications and was analyzed separately from the total number of PFA applications. A PFA application was considered completed when the energy delivery cycle was finished without system-triggered interruption. Data points were recorded and evaluated for every completed application, including instances in which a planned three-application set was terminated prematurely. Individual electrode pairs that were selectively deactivated because of catheter loop overlap or mutual contact, such as between electrodes 1 and 10, were excluded to ensure safe and appropriate pulse delivery. Consequently, 29,822 raw LI data points, comprising paired pre-ablation and post-ablation measurements, were evaluated from 1,556 total applications.

### Reproducibility of BiLID Measurement

To assess the reliability of video-based BiLID extraction, a random sample of 200 individual applications was independently reanalyzed by a second experienced electrophysiologist who was blinded to the original measurements and clinical outcomes. Interobserver agreement for extracted BiLID values was quantified using the intraclass correlation coefficient (ICC) with a two-way random-effects model for absolute agreement. An ICC >0.75 was considered to indicate good reproducibility, and an ICC >0.90 was considered to indicate excellent reproducibility.

### Blood Sampling and Biomarker Analysis

Venous blood samples were obtained following a previously described protocol.^9^To evaluate the acute and subacute biophysical effects of PFA, sampling was performed at three time points: T1, 24 to 72 hours before the procedure; T2, immediately after the final PFA application; and T3, the next morning on the first postoperative day. Myocardial response was assessed using creatine kinase-MB (CK-MB) and high-sensitivity cardiac troponin T (hs-cTnT). Serum haptoglobin and creatinine levels were also analyzed to monitor for intravascular hemolysis and renal impairment, respectively. For the analysis of myocardial response, peak biomarker levels obtained at T3 were used. These systemic biomarkers were treated as supportive, exploratory measures intended to provide biological corroboration of BiLID rather than as primary endpoints.

### Statistical Analysis

Continuous variables are expressed as mean ± standard deviation or median with interquartile range, as appropriate. Categorical variables are expressed as counts and percentages. Statistical significance was defined as a two-sided P value <0.05. All analyses were performed using JMP version 18.2.1 (SAS Institute, Cary, NC, USA). Analyses were conducted at two hierarchical levels. Repeated measurements obtained within patients, including individual energy applications and bipolar electrode-pair measurements, were analyzed using linear mixed-effects models. Patient-level summary parameters were analyzed using correlation and univariable regression analyses.

Longitudinal changes in biomarkers across the three time points were analyzed using repeated-measures analysis of variance, followed by post hoc pairwise comparisons against the baseline value (T1) and the immediate postprocedural value (T2).

Because multiple energy applications and bipolar electrode-pair measurements were obtained from each patient, application-level and electrode-pair-level analyses were performed using linear mixed-effects models to account for clustered repeated measurements. These models were estimated using restricted maximum likelihood, with patient identity included as a random intercept. Fixed effects were selected according to each comparison and included TPI status, the number of TPI-positive electrodes within a bipolar pair, application order within each three-application set, pulmonary vein side, and electrode pair. Pairwise comparisons of least-squares means were performed with Tukey adjustment when appropriate.

Correlations between patient-level procedural parameters, including Total BiLID, the number of applications, and the number of TPI-positive signals, and peak CK-MB levels were evaluated using Pearson correlation coefficients. Because of the relatively small patient sample size, univariable linear regression was used to evaluate clinical and procedural factors associated with peak CK-MB elevation at T3. Multivariable modeling was deliberately not performed to avoid overfitting, and the independence of individual patient-level factors was therefore not inferred. Categorical variables were coded as follows: sex, female = 1 and male = 0; and history of heart failure, yes = 1 and no = 0.

## Results

### Patient and Procedural Characteristics

Baseline clinical and procedural characteristics are summarized in Table 1. The study included 23 consecutive patients with a mean age of 65 ± 10 years; 14 patients (61%) were men. Paroxysmal atrial fibrillation was present in 17 patients (74%), and persistent atrial fibrillation was present in 6 patients (26%). A history of heart failure was noted in 7 patients (30%). The mean left atrial volume index was 33 ± 10 mL/m², and the median CHADS₂ score was 1 (interquartile range, 1–3).

**Table 1.** Patient Characteristics.

| Characteristics | N=23 |
| --- | --- |
| Age, years | 65 ± 10 |
| Male sex, n (%) | 14 (61) |
| Body mass index, kg/m<sup>2</sup> | 23.3 ± 2.7 |
| Paroxysmal atrial fibrillation, n (%) | 17 (74) |
| Persistent atrial fibrillation, n (%) | 6 (26) |
| History of heart failure, n (%) | 7 (30) |
| Hypertension, n (%) | 15 (65) |
| Diabetes mellitus, n (%) | 1 (4) |
| History of stroke, n (%) | 0 (0) |
| CHADS<sub>2</sub> score, median [IQR] | 1 [1–3] |
| LVEF, % | 63 ± 7 |
| LAD, mm | 37 ± 5 |
| LAVI, mL/m<sup>2</sup> | 33 ± 10 |
| Procedural Characteristics |  |
| Total PFA applications, n | 1,556 |
| PFA applications per patient, n | 67 ± 10 |
| Acute bidirectional PVI success, n (%) | 23 (100%) |
| Total procedure time, min | 68 ± 11 |
| Left atrial dwell time, min | 54 ± 16 |
| Total fluoroscopy time, min | 5.7 ± 3.2 |
| Major complications, n (%) | 0 (0%) |

Data are shown as mean ± SD, median [interquartile range], or n (%).
3 AF indicates atrial fibrillation; CHADS<sub>2</sub>, congestive heart failure, hypertension, age
4 ≥75 years, diabetes mellitus, and stroke; LAD, left atrial dimension; LAVI, left atrial
5 volume index; LVEF, left ventricular ejection fraction; PFA, pulsed-field ablation; and
PVI, pulmonary vein isolation.

A total of 1,556 completed PFA applications were analyzed offline, corresponding to 67 ± 10 applications per patient. From these applications, 29,822 individual local impedance data points, comprising paired pre-ablation and post-ablation measurements, were extracted and evaluated by frame-by-frame video analysis. Acute bidirectional PVI was achieved in all patients. The mean procedure time, left atrial dwell time, and fluoroscopy time were 68 ± 11 minutes, 54 ± 16 minutes, and 5.7 ± 3.2 minutes, respectively. No major periprocedural complications were observed.

### Reproducibility and Biophysical Behavior of BiLID

Interobserver reproducibility of video-based BiLID extraction was excellent. In the random sample of 200 independently reanalyzed applications, the intraclass correlation coefficient between the two blinded observers was 0.94 (95% CI, 0.91–0.98), indicating excellent agreement.

Representative biophysical responses and local impedance dynamics during PFA are shown in Figure 1. The distribution of unipolar TPI signals was displayed on the three-dimensional mapping interface (Figure 1A), while bipolar local impedance was monitored across adjacent electrode pairs on the variable-loop catheter (Figure 1B). PFA delivery produced an immediate decrease in bipolar local impedance from the pre-application baseline across actively coupling electrode pairs (Figure 1C). This paired pre-to-post application impedance drop, defined as BiLID, showed reproducible kinetic profiles across sequential applications delivered at the same target region (Figure 1D).

### BiLID and Contact Quality: A Graded Response Complementary to Binary TPI

We examined the relationship between binary TPI and continuous BiLID at the application and bipolar electrode-pair levels using linear mixed-effects models that accounted for clustered repeated measurements within patients. Electrode pairs associated with a positive TPI signal had higher BiLID values than TPI-negative pairs (P < 0.0001; Figure 2A).

**Figure 2.**
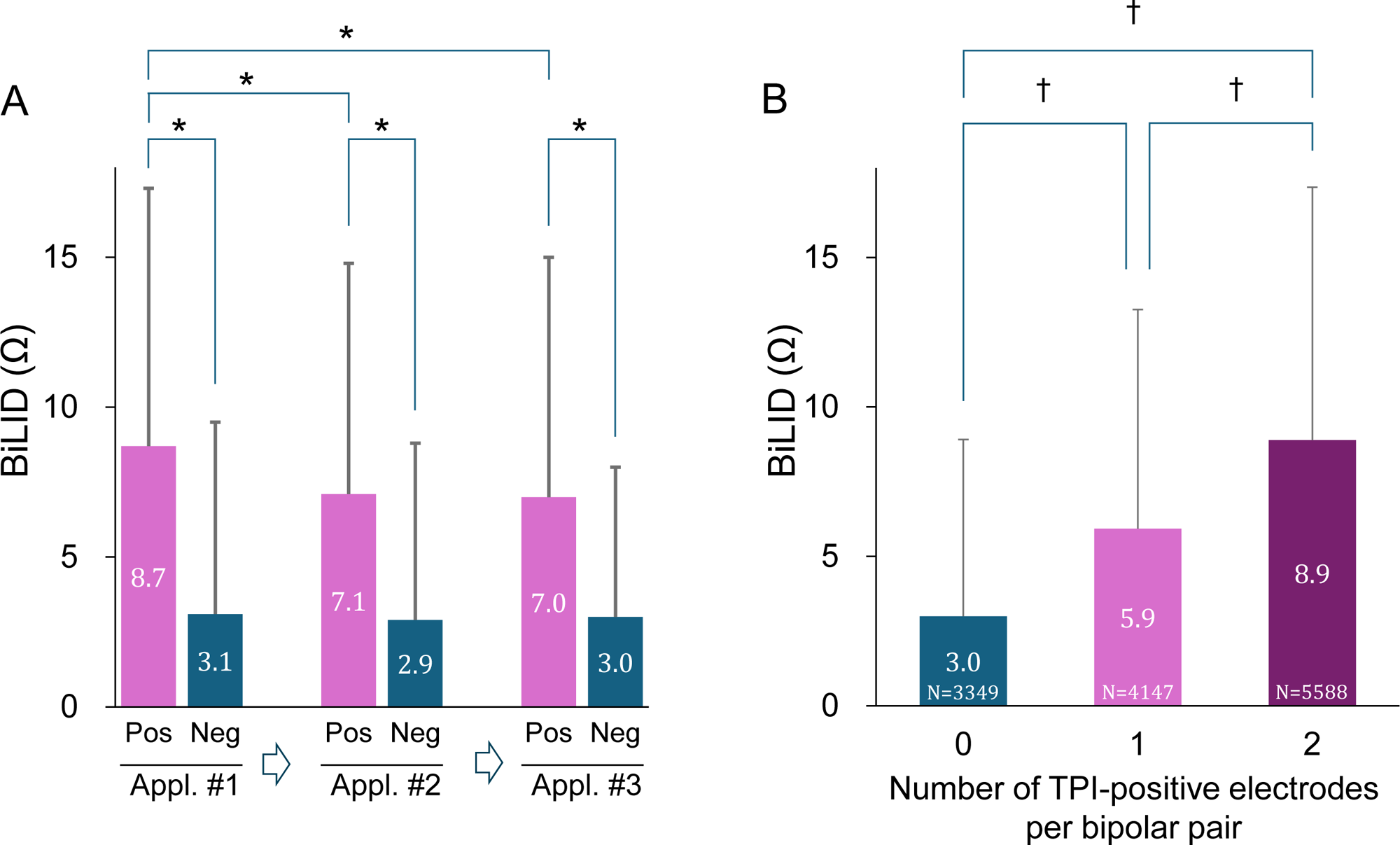
Relationship between the bipolar local impedance delta and tissue proximity indication. A, Comparison of BiLID between TPI-positive and TPI-negative electrode pairs according to application order within each three-application set. TPI-positive electrode pairs had higher BiLID values than TPI-negative pairs after accounting for clustered repeated measurements using linear mixed-effects models (P < 0.0001). Among TPI-positive electrode pairs, BiLID differed according to application order (mixed-effects model, P < 0.0001), with higher values during the first application than during the second and third applications. B, Effect of the number of TPI-positive electrodes within a bipolar pair on BiLID. BiLID increased stepwise as the number of TPI-positive electrodes rose from 0 to 1 to 2 (P < 0.0001). Asterisks indicate significant Tukey-adjusted pairwise differences in Figure 2A. Daggers indicate significant Tukey-adjusted pairwise differences among TPI-positive electrode-count groups in Figure 2B. Data are shown as mean ± SD. BiLID indicates Bipolar Local Impedance Delta; and TPI, tissue proximity indication.

Among TPI-positive electrode pairs, BiLID differed according to application order within each three-application set (mixed-effects model, P < 0.0001). Tukey-adjusted pairwise comparisons showed that BiLID was higher during the first application than during the second and third applications, whereas no difference was observed between the second and third applications.

BiLID also increased in a graded manner according to the number of TPI-positive electrodes within each bipolar pair. In the mixed-effects model, the number of TPI-positive electrodes was associated with BiLID, with a stepwise increase from 0 to 1 to 2 TPI-positive electrodes (P < 0.0001; Figure 2B). This relationship remained significant after accounting for clustered repeated measurements, indicating that BiLID captured quantitative variation in catheter–tissue coupling that was not represented by binary TPI alone.

### Spatial Heterogeneity of BiLID

Spatial heterogeneity of BiLID was evaluated using linear mixed-effects models accounting for clustered repeated measurements within patients. BiLID varied across loop electrode positions (P < 0.0001; Figure 3A). At the regional level, BiLID was lower during right pulmonary vein ablation than during left pulmonary vein ablation after adjustment for electrode position and within-patient clustering (P < 0.0001; Figure 3B). These findings indicate that both electrode position and pulmonary vein side influence the magnitude of catheter–tissue energy coupling.

**Figure 3.**
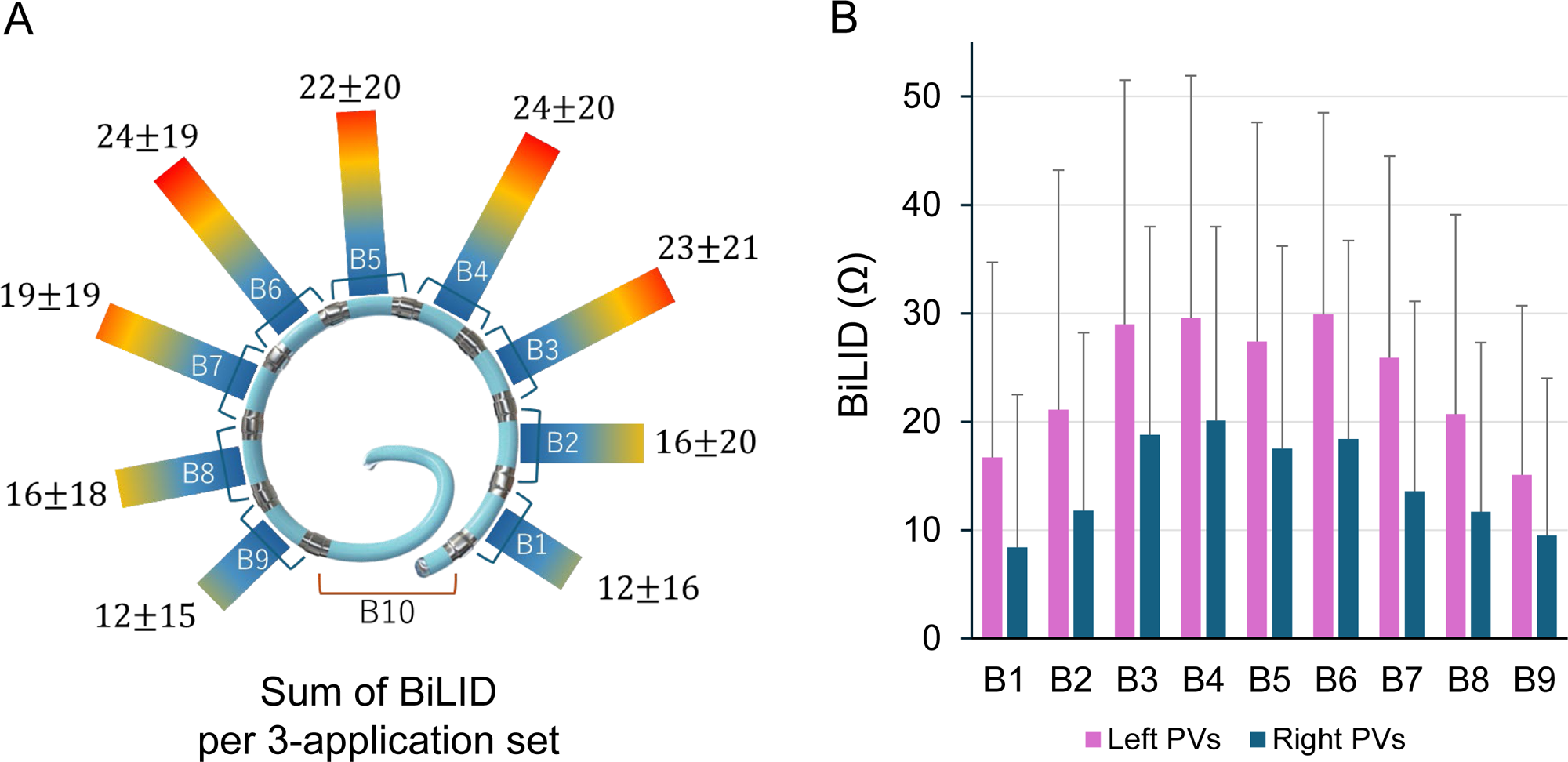
Regional distribution and comparison of BiLID across electrodes. A, Distribution of BiLID across loop electrode positions. The schematic illustrates the magnitude of BiLID at each bipolar electrode position on the variable-loop catheter. BiLID differed across electrode positions after accounting for clustered repeated measurements using a linear mixed-effects model (P < 0.0001). B, Comparison of BiLID between left and right pulmonary vein ablation. BiLID was lower during right pulmonary vein ablation than during left pulmonary vein ablation after adjustment for electrode position and within-patient clustering (P < 0.0001). Data are shown as mean ± SD. BiLID indicates Bipolar Local Impedance Delta; LPV, left pulmonary vein; and RPV, right pulmonary vein.

### Systemic Myocardial Response and Its Correlation with BiLID

Serum CK-MB and hs-cTnT levels increased after the procedure, with marked elevations at T2 and peak concentrations at T3 relative to baseline T1 values (all P < 0.05 on post hoc pairwise comparison; Table 2). Serum haptoglobin decreased at T3 (P < 0.05), consistent with transient procedure-related intravascular hemolysis, whereas serum creatinine and estimated glomerular filtration rate remained stable throughout the observation period, with no instances of acute kidney injury.

**Table 2.**
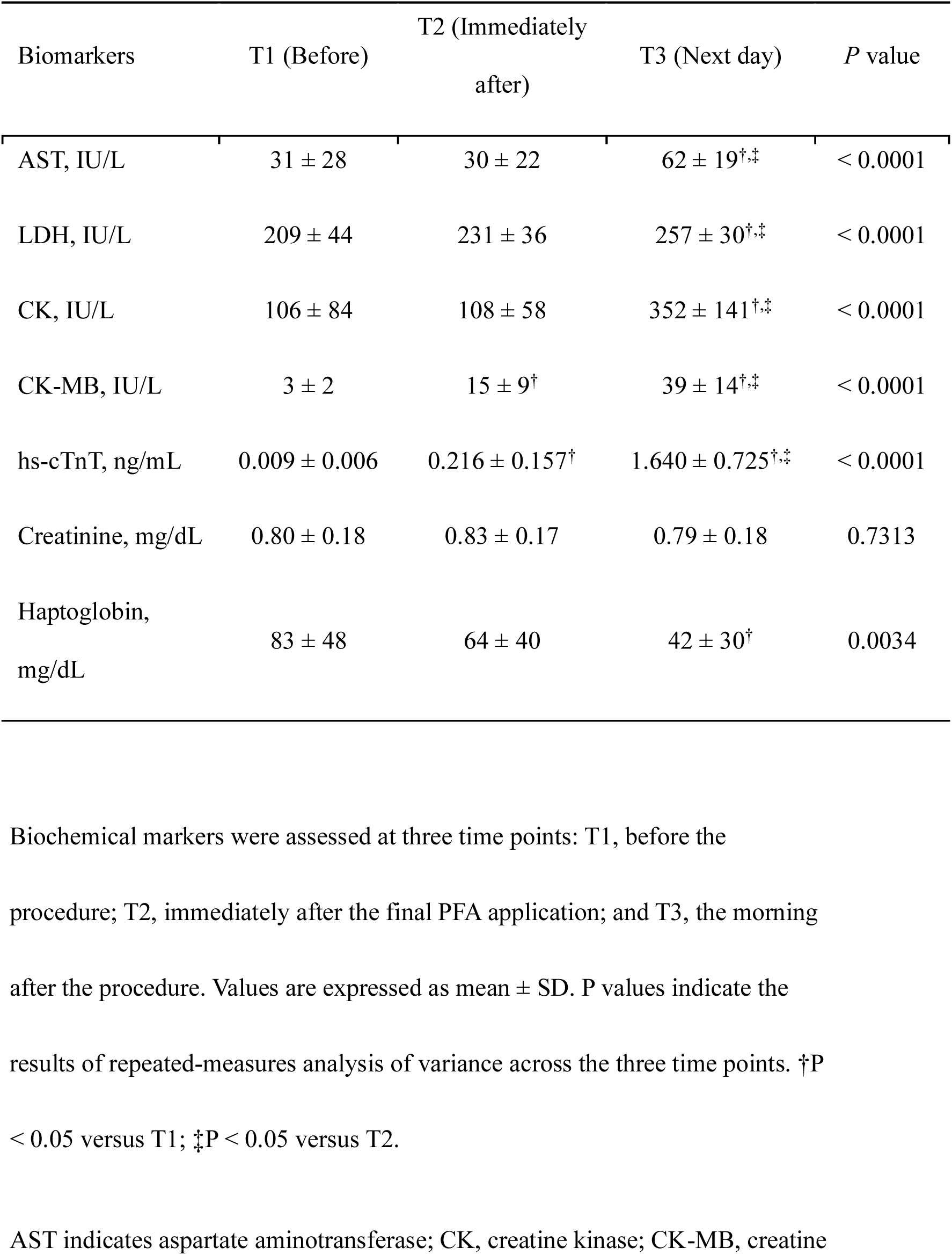
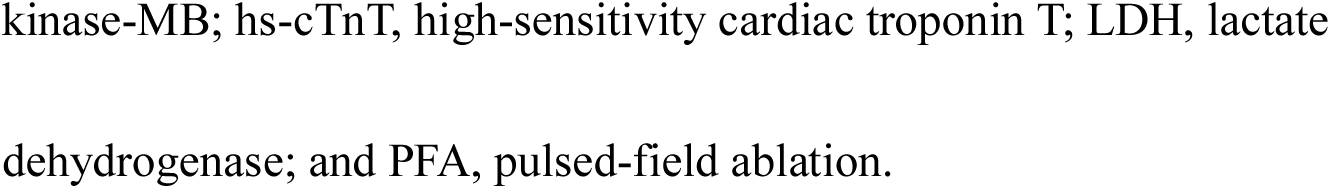
Longitudinal Changes in Biochemical Markers.

| Biomarkers | T1 (Before) | T2 (Immediately<br>after) | T3 (Next day) | *P* value |
| --- | --- | --- | --- | --- |
| AST, IU/L | 31 ± 28 | 30 ± 22 | 62 ± 19<sup>†,‡</sup> | < 0.0001 |
| LDH, IU/L | 209 ± 44 | 231 ± 36 | 257 ± 30<sup>†,‡</sup> | < 0.0001 |
| CK, IU/L | 106 ± 84 | 108 ± 58 | 352 ± 141<sup>†,‡</sup> | < 0.0001 |
| CK-MB, IU/L | 3 ± 2 | 15 ± 9<sup>†</sup> | 39 ± 14<sup>†,‡</sup> | < 0.0001 |
| hs-cTnT, ng/mL | 0.009 ± 0.006 | 0.216 ± 0.157<sup>†</sup> | 1.640 ± 0.725<sup>†,‡</sup> | < 0.0001 |
| Creatinine, mg/dL | 0.80 ± 0.18 | 0.83 ± 0.17 | 0.79 ± 0.18 | 0.7313 |
| Haptoglobin,<br>mg/dL | 83 ± 48 | 64 ± 40 | 42 ± 30<sup>†</sup> | 0.0034 |

3 Biochemical markers were assessed at three time points: T1, before the
4 procedure; T2, immediately after the final PFA application; and T3, the morning
5 after the procedure. Values are expressed as mean ± SD. *P* values indicate the
6 results of repeated-measures analysis of variance across the three time points. <sup>†</sup>*P*
7 < 0.05 versus T1; <sup>‡</sup>*P* < 0.05 versus T2.
8 AST indicates aspartate aminotransferase; CK, creatine kinase; CK-MB, creatine
- 1 kinase-MB; hs-cTnT, high-sensitivity cardiac troponin T; LDH, lactate - 2 dehydrogenase; and PFA, pulsed-field ablation. - 3

As biological corroboration of BiLID, we examined the relationship between patient-level procedural parameters and peak CK-MB at T3. Neither the total number of applications (r = 0.16, P = 0.4711; Figure 4A) nor the number of TPI-positive signals (r = 0.26, P = 0.2334; Figure 4B) correlated with peak CK-MB. In contrast, Total BiLID showed a strong positive correlation with peak CK-MB (r = 0.71, P = 0.0001; Figure 4C). Next-day hs-cTnT levels did not correlate with any of the evaluated procedural parameters (all P > 0.05).

**Figure 4.**
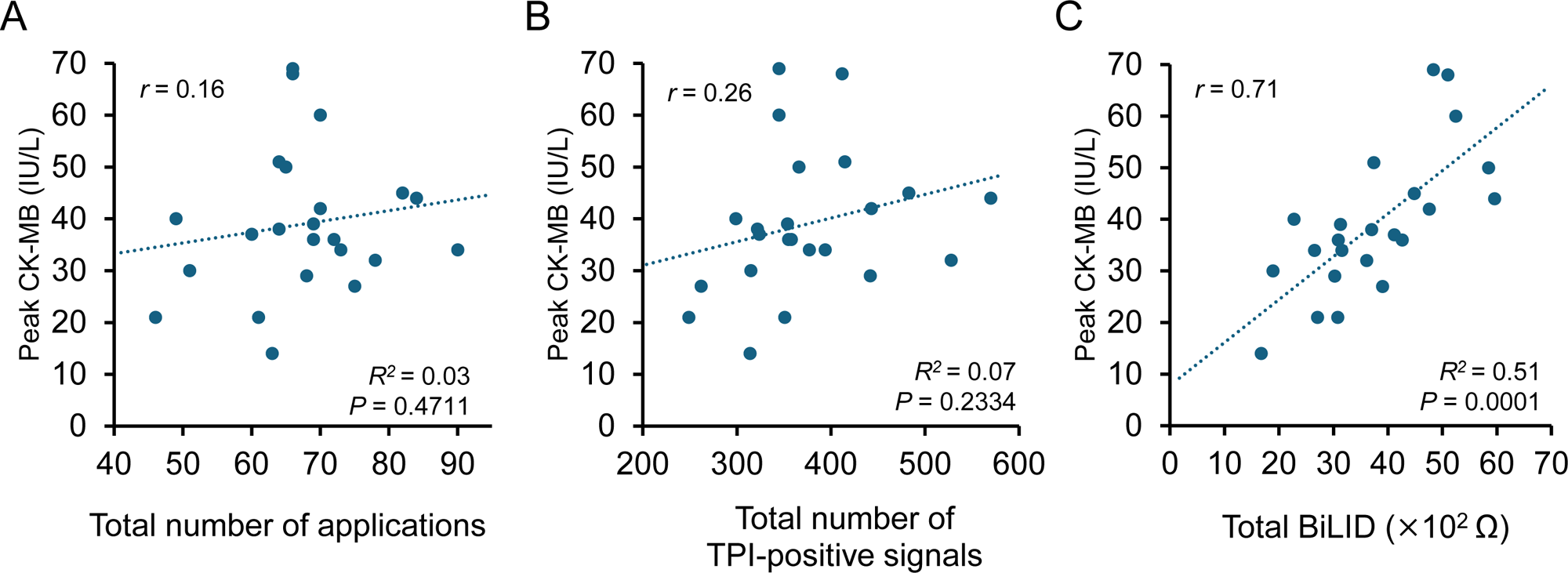
Correlations between procedural parameters and peak CK-MB levels. A, Relationship between the total number of applications and peak CK-MB levels (r = 0.16, R² = 0.025, P = 0.4711). B, Relationship between the total number of TPI-positive signals and peak CK-MB levels (r = 0.26, R² = 0.066, P = 0.2334). C, Relationship between Total BiLID and peak CK-MB levels (r = 0.71, R² = 0.505, P = 0.0001). Total BiLID, but not application count or TPI-positive signals, correlated with peak CK-MB. Dotted lines represent linear regression. P values and correlation coefficients were calculated using Pearson correlation analysis. BiLID indicates Bipolar Local Impedance Delta; CK-MB, creatine kinase-MB; and TPI, tissue proximity indication.

### Substrate-Related Factors Associated With Biomarker Release

Although Total BiLID was the strongest correlate of CK-MB release, patient-specific substrate factors were also associated with this biological response (Table 3). In univariable linear regression, left atrial volume index was inversely associated with peak CK-MB at T3 (standardized β = −0.57, P = 0.0045; Figure 5A). Lower CK-MB elevation was also associated with female sex (standardized β = −0.45, P = 0.0328; Figure 5B) and a history of heart failure (standardized β = −0.56, P = 0.0058; Figure 5C). These findings suggest that systemic biomarker release may be attenuated in more extensively remodeled atrial substrates, even when procedural energy coupling is quantified by BiLID.

**Figure 5.**
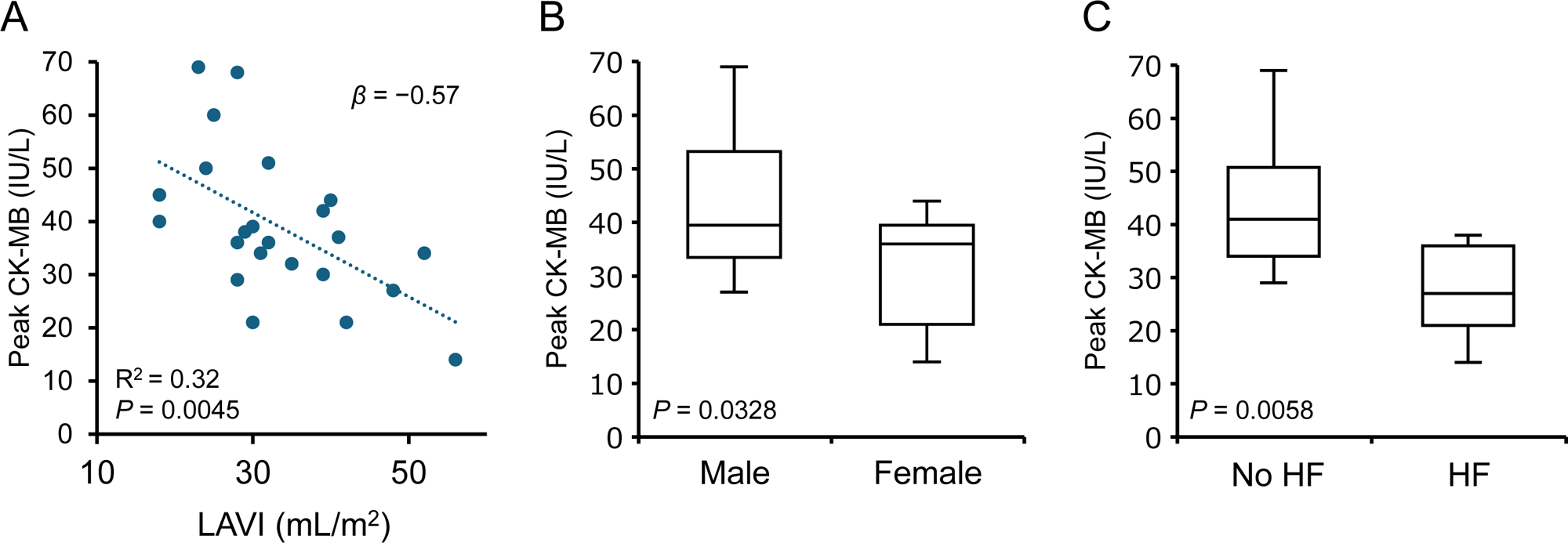
Substrate-related factors associated with peak creatine kinase-MB release. A, Relationship between left atrial volume index and peak CK-MB levels. A larger left atrial volume index was associated with lower peak CK-MB levels (standardized β = −0.57, R² = 0.324, P = 0.0045). The dotted line represents linear regression. B, Peak CK-MB levels according to sex. Women had lower peak CK-MB levels than men (P = 0.0328). C, Peak CK-MB levels according to history of heart failure. Patients with a history of heart failure had lower peak CK-MB levels than those without heart failure (P = 0.0058). In the box-and-whisker plots, the horizontal line within the box represents the median, the box represents the interquartile range, and the whiskers represent the 5th and 95th percentiles. P values were derived from univariable linear regression analyses, consistent with Table 3. CK-MB indicates creatine kinase-MB; HF, heart failure; and LAVI, left atrial volume index.

**Table 3.**
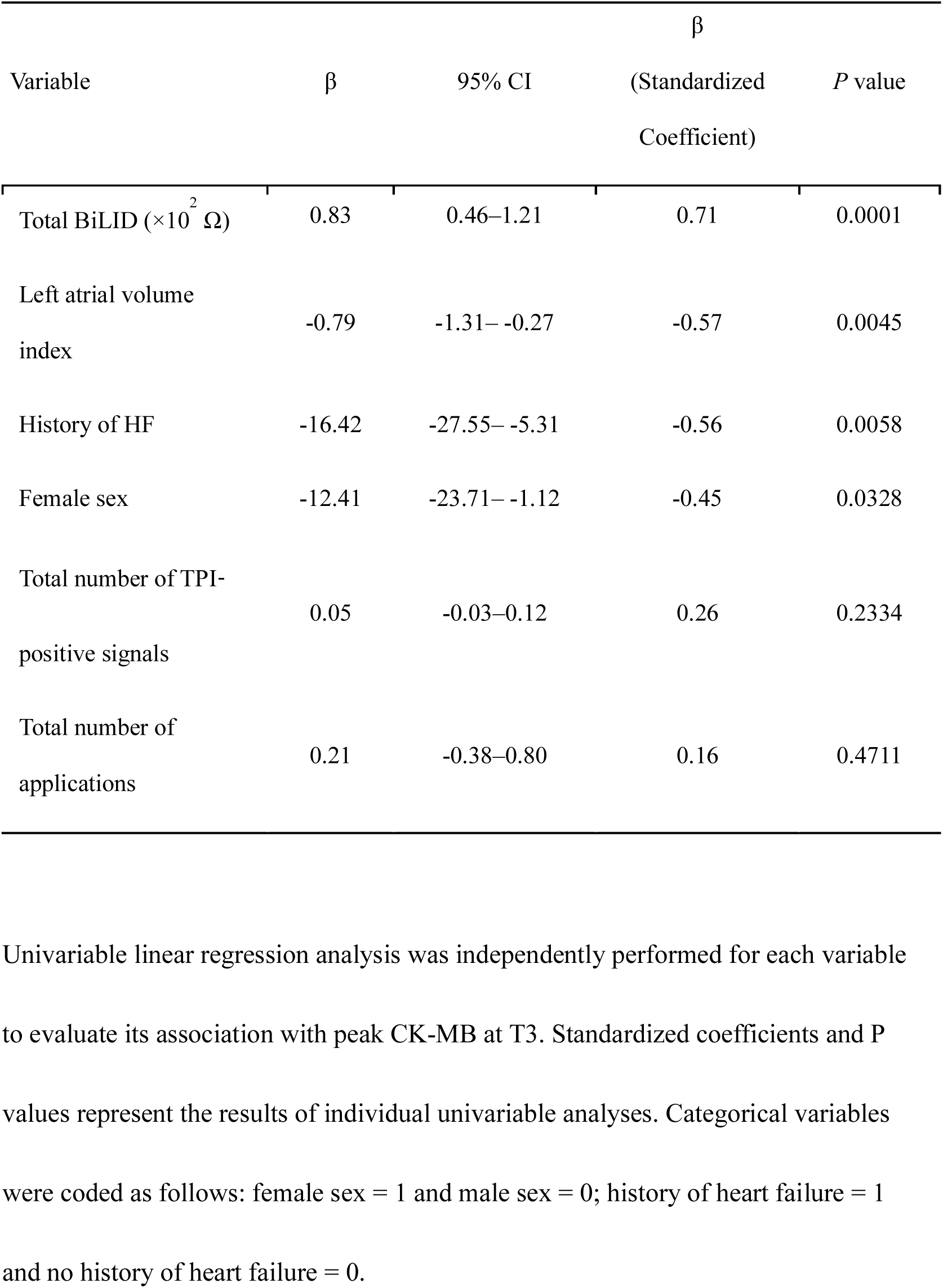
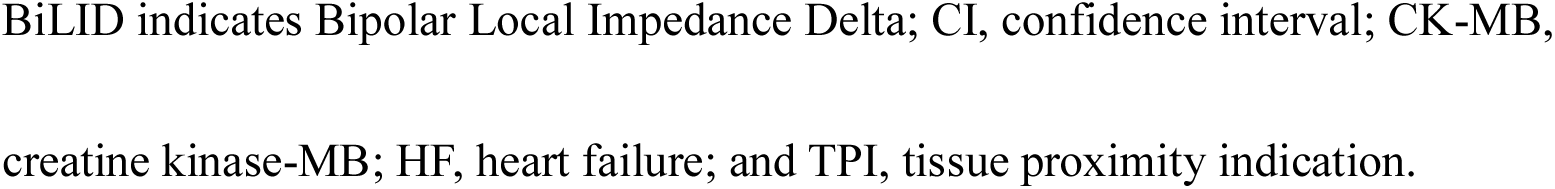
Univariable Linear Regression Analysis of Factors Associated With Peak CK-MB Release.

| Variable | $\beta$ | 95% CI | $\beta$<br>(Standardized<br>Coefficient) | *P* value |
| --- | --- | --- | --- | --- |
| Total BiLID ($\times 10^2 \Omega$) | 0.83 | 0.46–1.21 | 0.71 | 0.0001 |
| Left atrial volume<br>index | -0.79 | -1.31– -0.27 | -0.57 | 0.0045 |
| History of HF | -16.42 | -27.55– -5.31 | -0.56 | 0.0058 |
| Female sex | -12.41 | -23.71– -1.12 | -0.45 | 0.0328 |
| Total number of TPI-positive signals | 0.05 | -0.03–0.12 | 0.26 | 0.2334 |
| Total number of<br>applications | 0.21 | -0.38–0.80 | 0.16 | 0.4711 |

4 Univariable linear regression analysis was independently performed for each variable 5 to evaluate its association with peak CK-MB at T3. Standardized coefficients and *P* 6 values represent the results of individual univariable analyses. Categorical variables 7 were coded as follows: female sex = 1 and male sex = 0; history of heart failure = 1 8 and no history of heart failure = 0.
- 1 BiLID indicates Bipolar Local Impedance Delta; CI, confidence interval; CK-MB, - 2 creatine kinase-MB; HF, heart failure; and TPI, tissue proximity indication.

## Discussion

### Principal Findings

This study introduces Bipolar Local Impedance Delta (BiLID) as a continuous biophysical index derived from peri-application local impedance recordings during PFA using a variable-loop catheter system. Four principal findings emerged. First, video-based BiLID measurement showed excellent interobserver reproducibility. Second, BiLID increased in a graded manner with contact quality, rising stepwise with the number of TPI-positive electrodes within a bipolar pair, while also varying widely among TPI-positive signals. These findings indicate that BiLID captures information not represented by binary TPI alone. Third, BiLID exhibited spatial heterogeneity, with lower values during right pulmonary vein ablation than during left pulmonary vein ablation. Fourth, Total BiLID was biologically corroborated by a strong correlation with systemic CK-MB release, although biomarker release was also influenced by patient-specific substrate factors. Collectively, these findings support BiLID as a quantitative readout of catheter–tissue energy coupling that complements, rather than replaces, existing contact indicators.

### A Continuous Readout Complementary to Binary Tissue Proximity Indication

The central contribution of BiLID is that it adds a continuous, graded layer to an otherwise binary contact-assessment framework. TPI confirms that an electrode is in proximity to tissue, and the InspIRE experience established the clinical value of this information.^2^However, a positive TPI signal may be triggered by minimal or point-like contact, whereas a larger BiLID likely reflects a broader and more effective catheter–tissue interface that permits greater local energy coupling. Our observation that BiLID varied widely among TPI-positive signals directly quantifies this gap, and the stepwise increase in BiLID as the number of TPI-positive electrodes rose from 0 to 2 provides evidence of a graded relationship between contact quality and energy coupling.

BiLID therefore functions as a quantitative complement to TPI by linking simple proximity information with the magnitude of the local biophysical response to energy delivery. If implemented as an automated intraprocedural display, BiLID could help identify regions of relatively low coupling and provide a physiological basis for deciding when catheter repositioning or additional applications may be warranted.

### Biophysical Basis of BiLID

The acute impedance shift captured by BiLID likely reflects several concurrent physical and biological processes occurring at the catheter–tissue interface. Although PFA is commonly regarded as a nonthermal ablation modality, it remains an energy-transfer process in which high-voltage pulses can alter local tissue conductivity. Recent mathematical modeling by Farina et al.^10^introduced the concept of electrothermal coupling in PFA, showing that high-voltage pulse delivery can induce transient Joule heating. In addition, irrigation of PFA electrodes has been reported to mitigate Joule heating and heat-stacking phenomena, supporting the relevance of electrothermal effects at the catheter–tissue interface.^11^Even when clinically relevant thermal injury is not the intended mechanism of PFA, such transient temperature changes may increase ion mobility and tissue conductivity, thereby contributing to an acute decrease in local impedance. A second, complementary mechanism is the biological effect of the electric field on the cell membrane. In an intravascular PFA model, Wang et al.^12^demonstrated that electroporation-related disruption of the sarcolemma increases tissue conductivity as nanopores form and the previously insulating lipid bilayer becomes more conductive. BiLID may therefore represent a composite impedance response that reflects both conductivity changes related to transient electrothermal effects and membrane-level changes induced by electroporation.

This framework provides a plausible explanation for why BiLID tracked catheter–tissue energy coupling more closely than application counts or TPI-positive signals.

Application counts quantify how many energy deliveries occurred, and TPI confirms tissue proximity, but neither metric directly reflects the magnitude of the local impedance response to each delivery. By contrast, BiLID captures the immediate pre-to-post application change in bipolar local impedance, providing a clinically observable surrogate of local energy coupling. We emphasize, however, that BiLID remains an indirect marker of these underlying processes, and the relative contributions of electrothermal effects, membrane electroporation, catheter orientation, tissue apposition, and substrate characteristics cannot be resolved from the present clinical data.

### Spatial Heterogeneity and Catheter–Tissue Coupling

A notable finding of the present study was the spatial heterogeneity of BiLID, with lower values during right pulmonary vein ablation than during left pulmonary vein ablation. This regional difference likely reflects anatomical constraints and the resulting variation in catheter stability, loop orientation, and tissue apposition. In clinical practice, stable coaxial alignment of the variable-loop catheter is often more readily achieved in the left pulmonary veins, where the transseptal puncture site provides favorable backup support and a relatively direct vector for catheter advancement. In contrast, engagement of the right pulmonary veins frequently requires more complex sheath and catheter maneuvers, which may reduce the stability and surface area of catheter–tissue contact, particularly in posterior and inferior segments.

This regional pattern may have clinical relevance. Although the InspIRE trial established the overall efficacy of the variable-loop PFA system, detailed analyses of repeat procedures have identified right-sided posterior and inferior pulmonary vein segments as common sites of chronic reconnection.^2,7^ The lower BiLID values observed during right pulmonary vein ablation in the present study may provide a plausible biophysical explanation for this pattern, suggesting that reduced acute energy coupling in these anatomically challenging regions could contribute to subsequent lesion vulnerability.

From a mechanistic standpoint, even small gaps, unstable catheter orientation, or partial contact at the catheter–tissue interface may divert a greater proportion of current into the blood pool and reduce the local impedance response. BiLID may therefore help identify regions in which nominally adequate application delivery is accompanied by relatively low effective coupling. If incorporated into automated intraprocedural feedback systems, such information could guide catheter repositioning, contact optimization, or additional applications in anatomically vulnerable regions.

### Substrate as a Complementary Determinant of Biomarker Release

A further insight from this study is that BiLID and systemic biomarkers provide complementary, rather than interchangeable, information. BiLID quantifies the local impedance response associated with energy coupling at the time of delivery, whereas CK-MB reflects the systemic release of myocardial enzymes from tissue capable of responding to that delivery. In the present patient-level analysis, Total BiLID was the strongest procedural correlate of peak CK-MB release. However, substrate-related factors, including larger left atrial volume index, female sex, and a history of heart failure, were also associated with lower CK-MB elevation.

This pattern is best interpreted as a potentially source-limited biomarker response rather than as evidence of inadequate energy delivery. Chronic left atrial dilation is associated with structural remodeling, in which healthy atrial myocytes may be progressively replaced by interstitial fibrosis and fatty infiltration.^13^The pool of enzyme-rich viable myocytes available for electroporation may therefore be reduced, potentially attenuating systemic CK-MB release even when local energy coupling is quantified by BiLID. Consistent with this concept, Nakashima et al.^14^reported that women with atrial fibrillation often exhibit a greater burden of atrial fibrosis and lower bipolar voltage than men, suggesting reduced viable myocardial density. Chronic heart failure is also associated with structural and biochemical atrial remodeling that may reduce functional myocyte mass.^15,16^. These findings should not be interpreted as showing that BiLID is less valid in remodeled atria. Rather, they indicate that systemic biomarker release is likely determined by both the magnitude of local energy coupling and the amount of viable myocardial substrate available to respond. This distinction is important when interpreting biomarker-based assessments after PFA: a lower CK-MB response may reflect a more remodeled or source-limited substrate, not necessarily insufficient catheter–tissue coupling.

### Clinical Implications

These findings support BiLID as a quantitative complement to existing contact indicators in PFA. Current PFA workflows rely largely on cumulative application counts and binary proximity information, both of which are useful but insufficient to characterize the biophysical quality of each individual energy delivery. By providing a graded measure of catheter–tissue energy coupling, BiLID may help identify applications with relatively low effective coupling, recognize anatomically vulnerable regions such as right-sided pulmonary vein segments, and provide a physiological basis for catheter repositioning or additional applications.

The broader implication is a potential shift from application-count-based PFA toward coupling-guided lesion assessment. Although BiLID was derived offline in the present study, the index is based on peri-application local impedance changes that are already recorded and displayed during the procedure, requiring no additional measurement infrastructure. Future automated integration of BiLID into intraprocedural display systems may enable more individualized PFA titration and improve procedural reproducibility. A key practical advantage of BiLID is that it requires no proprietary data export or manufacturer support: it is computed entirely from the standard local impedance values already displayed on the mapping interface. This makes the index broadly reproducible across centers using the same system, without additional hardware or vendor-dependent log retrieval—an explicit design goal of the present study.

### Study Limitations

Several limitations should be acknowledged. First, this was a single-center retrospective study with a relatively small patient sample. Although the application-level and electrode-pair-level analyses were based on a large number of repeated measurements, patient-level associations, particularly those involving biomarkers and substrate-related factors, should be regarded as exploratory and require validation in larger multicenter cohorts.

Second, this study was designed as a biophysical proof-of-concept study and did not assess long-term clinical outcomes. The relationship between procedural BiLID and durable pulmonary vein isolation, chronic reconnection, or freedom from atrial tachyarrhythmia recurrence remains to be established. Third, BiLID was calculated offline from recorded procedural videos and was not available to the operator during the procedure. Therefore, the feasibility, accuracy, and clinical usefulness of real-time automated BiLID display require future technical development and prospective validation.

Fourth, BiLID is an indirect surrogate for catheter–tissue energy coupling.

Although the mechanistic interpretation was supported by prior modeling and experimental studies, direct intramyocardial temperature, electric field, and lesion-depth measurements were not available in this clinical setting. Fifth, BiLID was derived from a specific temporal window immediately before and after each energy delivery. Whether alternative sampling intervals or post-ablation impedance recovery kinetics provide additional predictive value remains unknown.

Finally, systemic biomarkers were used only as supportive biological corroboration. CK-MB release may reflect not only local energy coupling but also the amount of viable myocardial substrate, systemic biomarker kinetics, and patient-specific remodeling. Because of the limited sample size, multivariable modeling was not performed, and the independence of individual patient-level factors could not be formally established.

## Conclusion

This study introduces BiLID as a reproducible, continuous index of catheter–tissue energy coupling during PFA with a variable-loop catheter system. Unlike binary proximity indicators and cumulative application counts, BiLID provided a graded measure of energy coupling that varied with contact quality and pulmonary vein anatomy. Total BiLID correlated with systemic CK-MB release, whereas application count and TPI-positive signals did not, supporting its role as a biologically corroborated marker of effective coupling. These proof-of-concept findings suggest that BiLID may provide a framework for future individualized, coupling-guided PFA titration, warranting validation in larger prospective studies with long-term clinical outcomes.

## Data Availability

The data that support the findings of this study are available from the corresponding author upon reasonable request.

## Acknowledgements

None.

## Declaration of generative AI

The authors used ChatGPT (OpenAI) during manuscript preparation for language editing and clarity improvement. The tool was not used for data generation, statistical analysis, figure preparation, or scientific interpretation. All content was reviewed and verified by the authors, who take full responsibility for the final manuscript.

## Conflict of interest statement

- M. Kimura is an associate professor in the Department of Advanced Management of Cardiac Arrhythmias, an endowment department supported by Medtronic Japan Co., Ltd. and Japan Lifeline Co., Ltd. He has also received speaker honoraria from Medtronic Japan Co., Ltd.; Boston Scientific Japan Co., Ltd.; Johnson & Johnson; and Toray.
- M. Hiyama is affiliated with the Department of Cardiac Remote Management System, an endowment department supported by BIOTRONIK Japan Co., Ltd.
- S. Hamaura is affiliated with the Department of Advanced Therapeutics for Cardiovascular Diseases, an endowment department supported by Boston Scientific Japan Co., Ltd.
- S. Sasaki has received a research grant from Boston Scientific Japan Co., Ltd. and is a concurrent associate professor in the Department of Advanced Management of Cardiac Arrhythmias and the Department of Cardiac Remote Management System.
- H. Tomita has received a research grant from Abbott Medical Japan, LLC and is a concurrent professor in the Department of Advanced Management of Cardiac Arrhythmias, Department of Cardiac Remote Management System, and Department of Advanced Therapeutics for Cardiovascular Diseases.
- The other authors have no relevant disclosures.

None of the commercial entities listed above, including the manufacturer of the studied ablation system (Biosense Webster / Johnson & Johnson), had any role in the conception, design, conduct, analysis, or reporting of this investigator-initiated study.

## Nonstandard Abbreviations and Acronyms

AF =: atrial fibrillation
BiLID =: Bipolar Local Impedance Delta
CK-MB =: creatine kinase-MB
HF =: heart failure
hs-cTnT =: high-sensitivity cardiac troponin T
LA =: left atrium
LAVI =: left atrial volume index
LI =: local impedance
PFA =: pulsed-field ablation
PV =: pulmonary vein
PVI =: pulmonary vein isolation
TPI =: tissue proximity indication

---

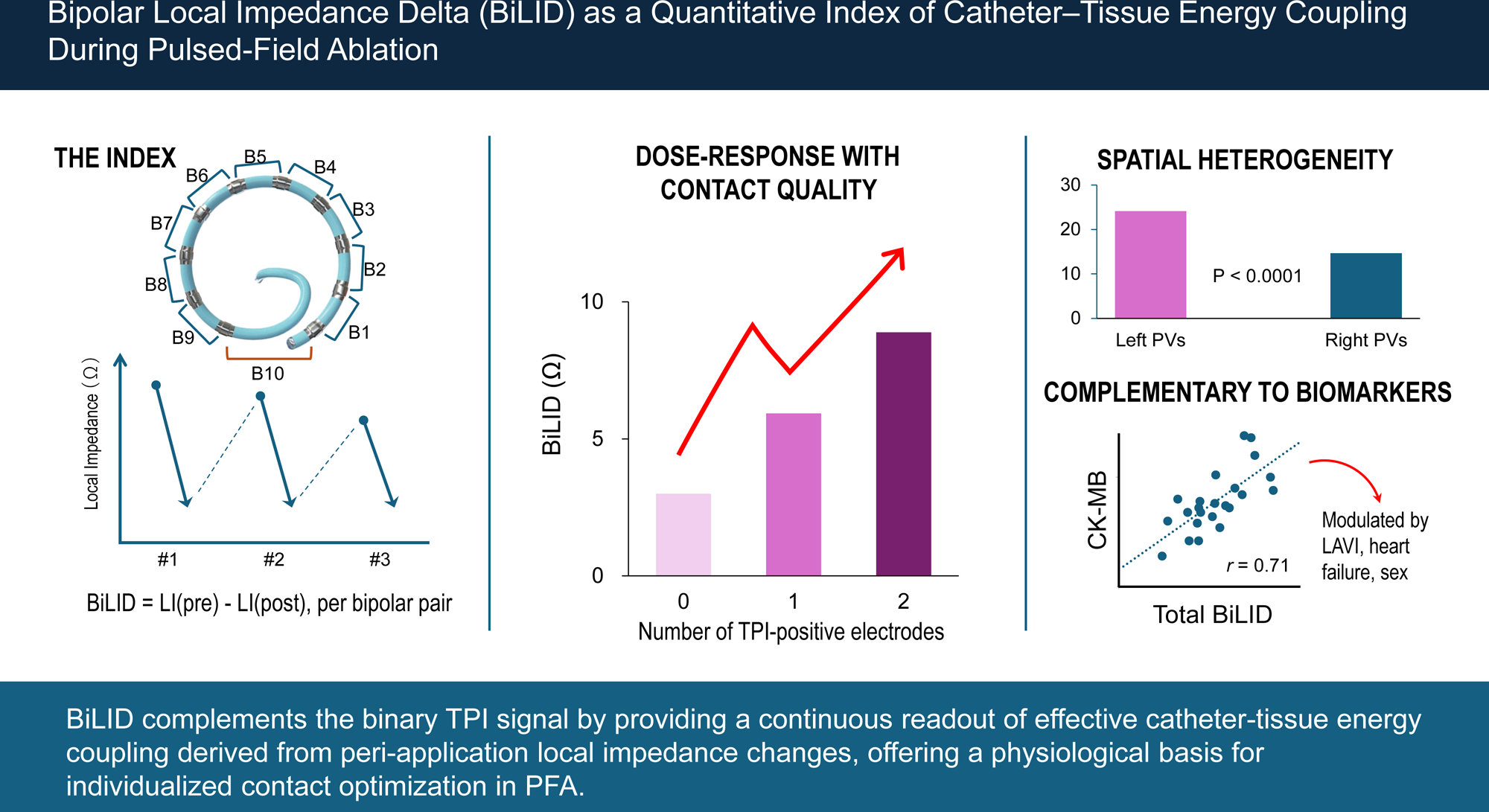

